# Nasal Mucosal Immunity is Associated with Protection against RSV Illness in Infants

**DOI:** 10.64898/2026.10.22.26366243

**Authors:** Faith B. Taliaferro, Ying Tang, Sergio Monares Ortiz, Olyvia J Jasset, Caroline Bald, Alison Araten, Calvin Fairbourn, Wanda I. Gonzalez, Peter P. Moschovis, Kathryn E. Stephenson, Andrea G. Edlow, Bruce H. Horwitz, Lael M. Yonker

**Affiliations:** Department of Pediatrics, Massachusetts General Hospital, Boston, MA; Division of Gastroenterology, Hepatology, and Nutrition, Boston Children’s Hospital, Boston, MA; Harvard Medical School, Boston, MA; Division of Pulmonary and Critical Care Medicine, Children’s Health, Dallas, TX; Department of Pediatrics, University of Texas Southwestern Medical Center, Dallas, TX; Center for Virology and Vaccine Research, Beth Israel Deaconess Medical Center, Boston, MA; Department of Obstetrics and Gynecology, Massachusetts General Hospital, Boston, MA; Division of Emergency Medicine, Boston Children’s Hospital, Boston, MA; Division of Pulmonary and Sleep Medicine, Children’s Health, Dallas, TX

**Keywords:** RSV, mucosal IgA, mucosal IgG, serological IgA, serological IgG, infants

## Abstract

**Background:** Despite advances in respiratory syncytial virus (RSV) immunoprophylaxis, RSV remains a leading cause of severe respiratory illness in infants. Immunity at the site of infection is potentially important yet airway mucosal immunity against RSV infection remains poorly defined.

**Methods:** We collected nasal and blood samples across three pediatric cohorts: 1) children presenting to the emergency department with respiratory symptoms; 2) infants and mothers presenting to an urban hospital system across the RSV season; 3) infants born to mothers vaccinated with maternal pre-Fusion (F) vaccine. We analyzed mucosal and serologic anti-RSV antibodies and assessed for factors impacting titer magnitude and durability.

**Results:** Specimens from 285 infants/young children and 39 mothers were analyzed. Mucosal anti-RSV antibody levels were lower in infants presenting with RSV infection compared to age-matched controls both in infants experiencing their first viral season (p = 0.0011) and older children (p < 0.0001). Higher levels of mucosal antibodies were seen in infants whose mothers received maternal pre-F vaccination, with IgG against RSV fusion protein tightly correlating between infant’s serum and nasal compartments (p < 0.0001). However, mucosal anti-RSV IgG waned significantly by 6 months of age (p < 0.0001).

**Conclusions:** Higher mucosal anti-RSV antibody levels are associated with protection against RSV-related illness. Current immunoprophylactic strategies drive robust anti-RSV IgG responses in the infant mucosal and serological compartments; however, passive immunity wanes over 6 months. These results suggest that RSV immunization strategies optimizing durable mucosal immunity may protect infants from RSV-related respiratory illness.

## Introduction

Respiratory syncytial virus (RSV) is a common cause of lower respiratory tract infections among infants and young children worldwide[1, 2], with the majority of children acquiring the virus by two years of age[3, 4]. Fortunately, two recent immunoprophylactic interventions, the maternal RSV pre-Fusion vaccine and the infant RSV neutralizing monoclonal antibody treatment nirsevimab, have both been shown to be effective in reducing the incidence of severe RSV-related illness in infants[5, 6]. However, surveillance reporting reveals that RSV remains a major cause of respiratory illness in young children[7], suggesting that optimized RSV prevention strategies and delivery remain essential.

While infant serological antibody responses related to RSV have been well characterized, mucosal immunity and its role in preventing the acquisition of RSV and/or limiting disease severity are less well understood. Given that the airway mucosal immune response represents one of the first lines of defense against inhaled pathogens, characterizing upper airway mucosal anti-RSV antibodies and understanding their relationship to protection is integral to the development of optimal protective strategies.

Here, we sought to determine whether nasal mucosal antibodies were associated with protection against RSV, and if so, what factors impact quantity and durability of mucosal antibody responses in infants. To achieve this, we analyzed nasal and/or blood samples from 285 infants/children and 39 mothers across three pediatric/infant cohorts: 1) children presenting to the emergency department with symptoms consistent with acute upper respiratory infections; 2) a cross-sectional cohort of infants and their mothers in an urban hospital system during an RSV season; 3) a longitudinal cohort of infants born to mothers who received the maternal pre-F vaccine during pregnancy (**Figure 1**). Our results illustrate the potential importance of upper airway mucosal antibodies in protection against RSV while highlighting factors that optimize anti-RSV nasal mucosal antibody levels in infants.

**Figure 1.**
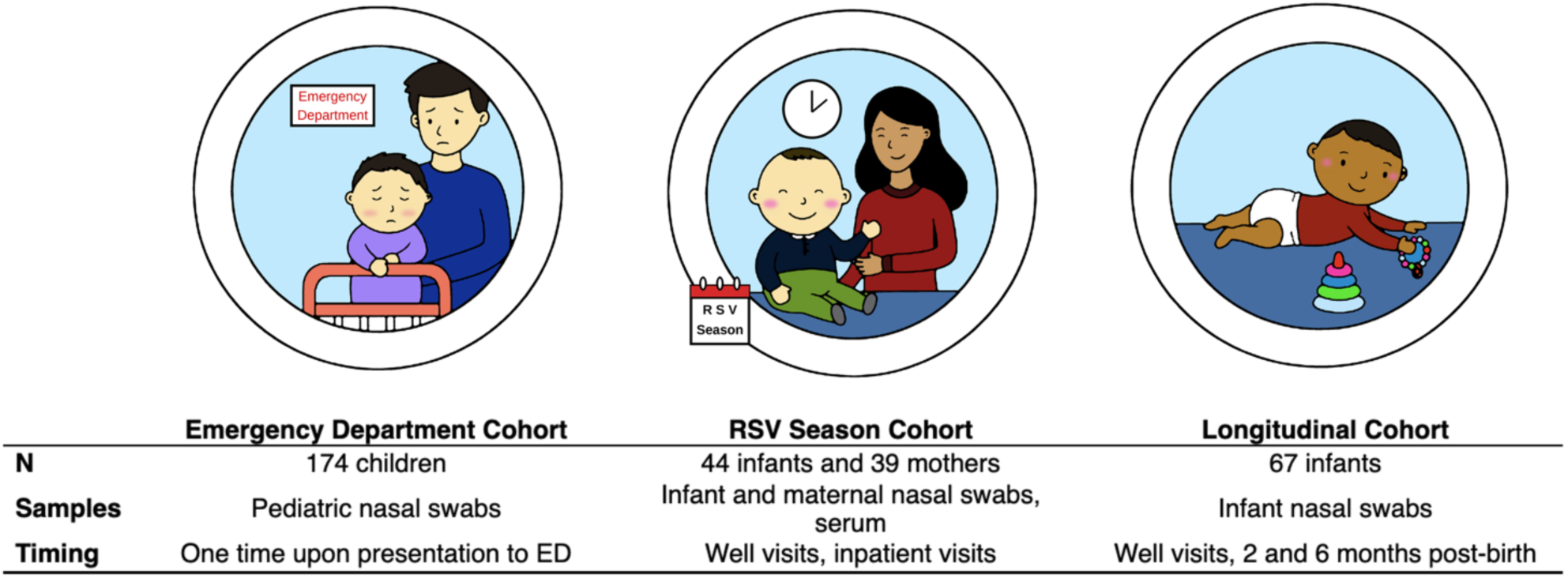
Cohort study design.

## Results

### Diminished nasal antibody levels are associated with increased likelihood of developing medically-attended RSV

To explore whether nasal antibody levels were associated with protection from medically attended RSV-related illness, we collected and analyzed nasal swabs from a convenience sample of children presenting to the Boston Children’s Hospital Emergency Department (ED) (“Emergency Department Cohort”) with symptoms consistent with respiratory infections across the three respiratory viral seasons (September through April: 2023-2024; 2024-2025; and 2025-2026). Participants were grouped by age and whether RSV was detected at time of presentation. Infants younger than 8 months of age were considered as likely experiencing their first viral season. One hundred and seventy-four children were included in the cohort. Mean participant age of this ED cohort was 19.7 months (standard deviation (SD) = 17.3 months). Of these children, 52 (30%) were ≤ 8 months of age, 61 (35%) were female, and 85 (49%) tested positive for RSV upon presentation (**Table 1**).

**Table 1.** Cohort demographics at time of enrollment.

|  | Cohort |  |  |  |
| --- | --- | --- | --- | --- |
|  | Overall<br>N = 285<br>(100%) | Emergency<br>Department<br>Cohort<br>N = 174<br>(61%) | RSV Season<br>Cohort<br>N = 44<br>(15%) | Longitudinal<br>Cohort<br>N = 67<br>(24%) |
| **Age at Enrollment (months)** |  |  |  |  |
| Mean (SD) | 12.8 (16.2) | 19.7 (17.3) | 4.6 (3.7) | 0.0 (0.0) |
| Median | 7.0 | 13.3 | 3.9 | 0.0 |
| Min, Max | 0.0, 80.8 | 1.6, 80.8 | 0.7, 17.2 | 0.0, 0.0 |
| **Sex** |  |  |  |  |
| Female | 112 (39%) | 61 (35%) | 20 (45%) | 31 (46%) |
| Male | 173 (61%) | 113 (65%) | 24 (55%) | 36 (54%) |
| **Race** |  |  |  |  |
| Asian | 14 (4.9%) | 9 (5.2%) | 0 (0%) | 5 (7.5%) |
| Black/African-American | 36 (13%) | 34 (20%) | 0 (0%) | 2 (3.0%) |
| White | 102 (36%) | 46 (26%) | 10 (23%) | 46 (69%) |
| Multiracial | 2 (0.7%) | 0 (0%) | 2 (4.5%) | 0 (0%) |
| Other | 44 (15%) | 31 (18%) | 0 (0%) | 13 (19%) |
| Declined/Not Listed | 87 (31%) | 54 (31%) | 32 (73%) | 1 (1.5%) |
| **Ethnicity** |  |  |  |  |
| Hispanic or Latino | 72 (25%) | 43 (25%) | 23 (52%) | 6 (9.0%) |
| Not Hispanic or Latino | 139 (49%) | 76 (44%) | 5 (11%) | 58 (87%) |
| Declined/Not Listed | 74 (26%) | 55 (32%) | 16 (36%) | 3 (4.5%) |

First, we examined whether nasal antibodies against the RSV fusion (F) protein were inversely associated with RSV positivity. Across the three-year study period, infants presenting with RSV illness during their first viral season had significantly lower nasal anti-RSV-F IgG levels compared to age-matched RSV-negative infants (**Figure 2A, left**, p = 0.0011). This suggests that higher levels of anti-RSV-F IgG in the mucosal compartment may be protective against RSV, although causality cannot be determined in this cross-sectional cohort. In contrast, nasal anti-RSV-F IgA levels did not differ significantly between infants ≤8 months of age presenting with RSV as compared to those without RSV (**Figure 2A, right**). Unlike IgG, IgA is not transferred transplacentally, nor is it a component of nirsevimab. Rather mucosal IgA is primarily induced following mucosal infection[8].

**Figure 2.**
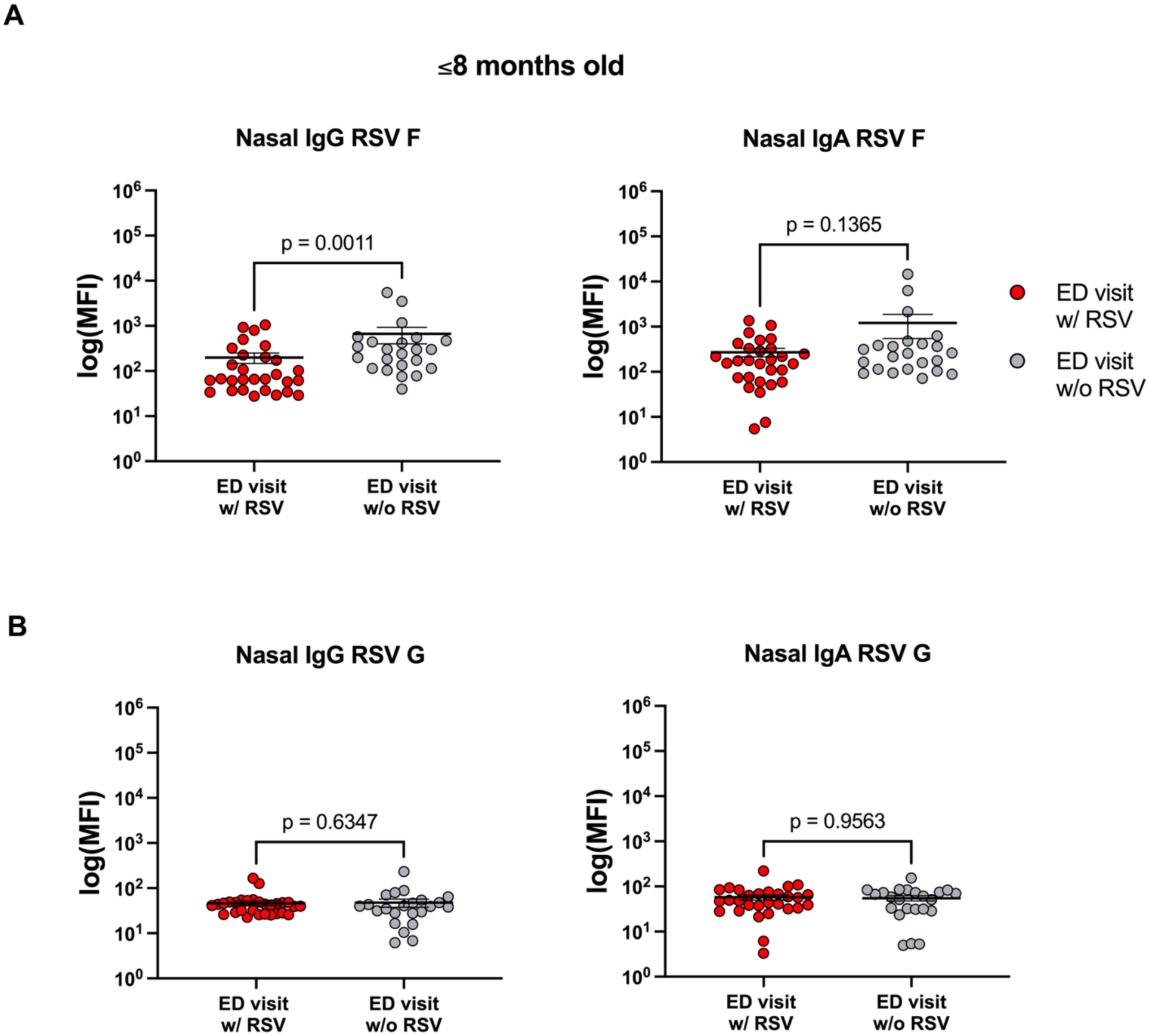
Nasal antibody titers in infants ≤8 months of age. Nasal IgG (left) and IgA (right) levels against RSV-A fusion (F) protein (**A**) and attachment (G) protein (**B**) in infants ≤8 months of age with RSV (n=29) and without RSV (n=23) are shown. Mean value and standard error of the mean are displayed. Two-tailed Mann-Whitney U tests were performed between individual groups. Abbreviations: Ig, immunoglobulin; MFI, median fluorescence intensity.

Maternal breast milk may also contribute to nasal IgA in infants, although this has not been defined. Additionally, breastfeeding histories were unavailable.

In this cohort of infants ≤8 months of age, nasal IgG and IgA against the RSV attachment (G) protein were uniformly low (**Figure 2B**), suggesting that differences in anti-RSV-F IgG were not driven by prior natural infection with RSV. Rather, this suggests that differences in anti- RSV-F IgG were due to passive exposures, either through maternal vaccination or infant nirsevimab, although we do not have vaccine history for this cohort. Similar patterns were observed for antibodies against RSV-B F and G proteins (**Supplemental Figure 1**). These data support a potential protective role for nasal anti-RSV-F IgG in enhancing infant protection against RSV.

We next evaluated children older than 8 months of age, presumed to have experienced at least one respiratory viral season, who presented to the emergency department with symptoms of respiratory infection. As in the younger cohort, RSV-positive children had significantly lower nasal anti-RSV-F IgG levels than RSV-negative children (**Figure 3A, left**, p < 0.0001; **Supplemental Figure 2A, left**). However, unlike the younger cohort, RSV-positive children also had significantly lower nasal anti-RSV-F IgA levels (**Figure 3A, right**, p < 0.0001; **Supplemental Figure 2A, right**), as well as lower nasal IgG and IgA levels against the RSV attachment (G) protein (**Figure 3B; Supplemental Figure 2B**). Multivariate analysis indicated that increases in both mucosal anti-RSV-F IgG and IgA were independently associated with lower odds ratio of infection with RSV (OR 0.50 and 0.51, respectively).

**Figure 3.**
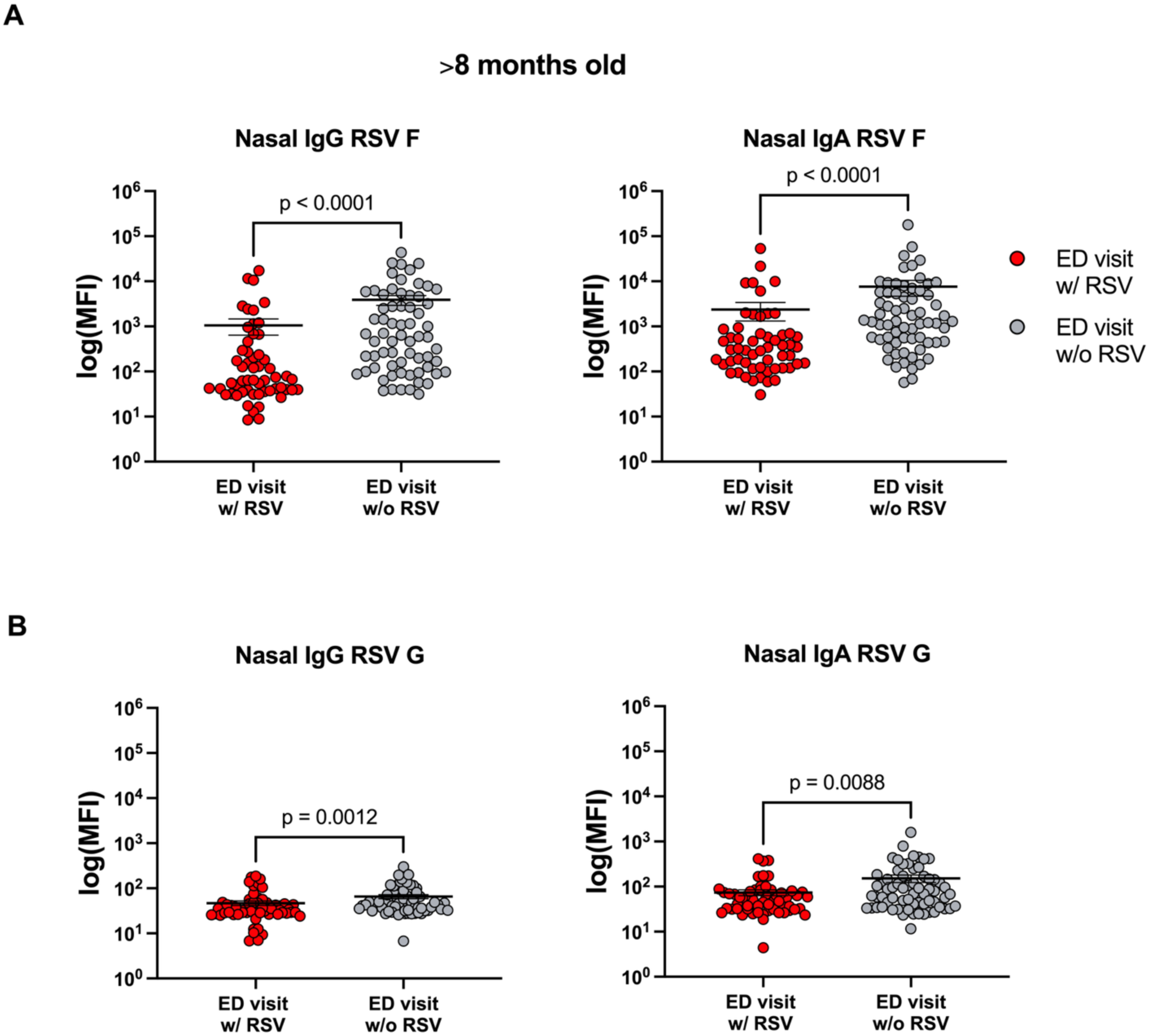
Nasal antibody titers in infants older than 8 months of age. Nasal IgG (left) and IgA (right) levels against RSV-A fusion (F) protein (**A**) and attachment (G) protein (**B**) in children >8 months of age with RSV (n=56) and without RSV (n=66) are shown are shown. Mean value and standard error of the mean are displayed. Two-tailed Mann-Whitney U tests were performed between individual groups. Abbreviations: Ig, immunoglobulin; MFI, median fluorescence intensity.

These findings suggest that prior natural RSV infection was more common among RSV-negative than RSV-positive children and support a potential protective role for both nasal IgG and IgA against subsequent RSV infection.

### Maternal pre-F vaccination contributes to infant upper airway immunity against RSV

To test whether maternal vaccination directly contributes to airway immunity against RSV, we analyzed blood and nasal samples collected from infants with well-characterized maternal vaccination histories (“RSV Season Cohort”; **Figure 1A**). Infant participants were grouped into three categories: 1) infants whose mothers received the maternal pre-F vaccine during pregnancy (“maternal pre-F”), 2) infants who received the monoclonal antibody nirsevimab (“nirsevimab”), and 3) infants who received no RSV immunoprophylaxis (“no RSV immunoprophylaxis”). Forty-four infants and their mothers were included in this cohort, with a mean infant age of 4.6 months (SD = 3.7 months) at time of enrollment, and 20 (45%) of the infants were female (**Figure 1B**).

Infants whose mothers received the pre-F vaccine during pregnancy had significantly higher serum and nasal levels of anti-RSV-F IgG compared to infants whose mothers did not receive any RSV immunoprophylaxis (**Figure 4A, left**, p = 0.0007; **Figure 4A, right**, p = 0.0022). Similar associations are seen for RSV-B (**Supplemental Figure 3A**). These results indicate that in addition to increasing infant levels of anti-RSV-F antibodies in the serum maternal immunization also elevates infant levels of anti-RSV-F antibodies in the nasal mucosa. This difference was not detected in infants who received nirsevimab as compared to no RSV immunoprophylaxis **(Supplemental Figure 4)**; differences in IgG RSV F transfer across mucosal surfaces warrants further study. As in the younger ED cohort, we did not see any differences seen in anti-RSV-A or anti-RSV-B nasal IgA levels **(Supplemental Figures 5A, 6A)**, suggesting that these infants had not previously experienced natural infection.

**Figure 4.**
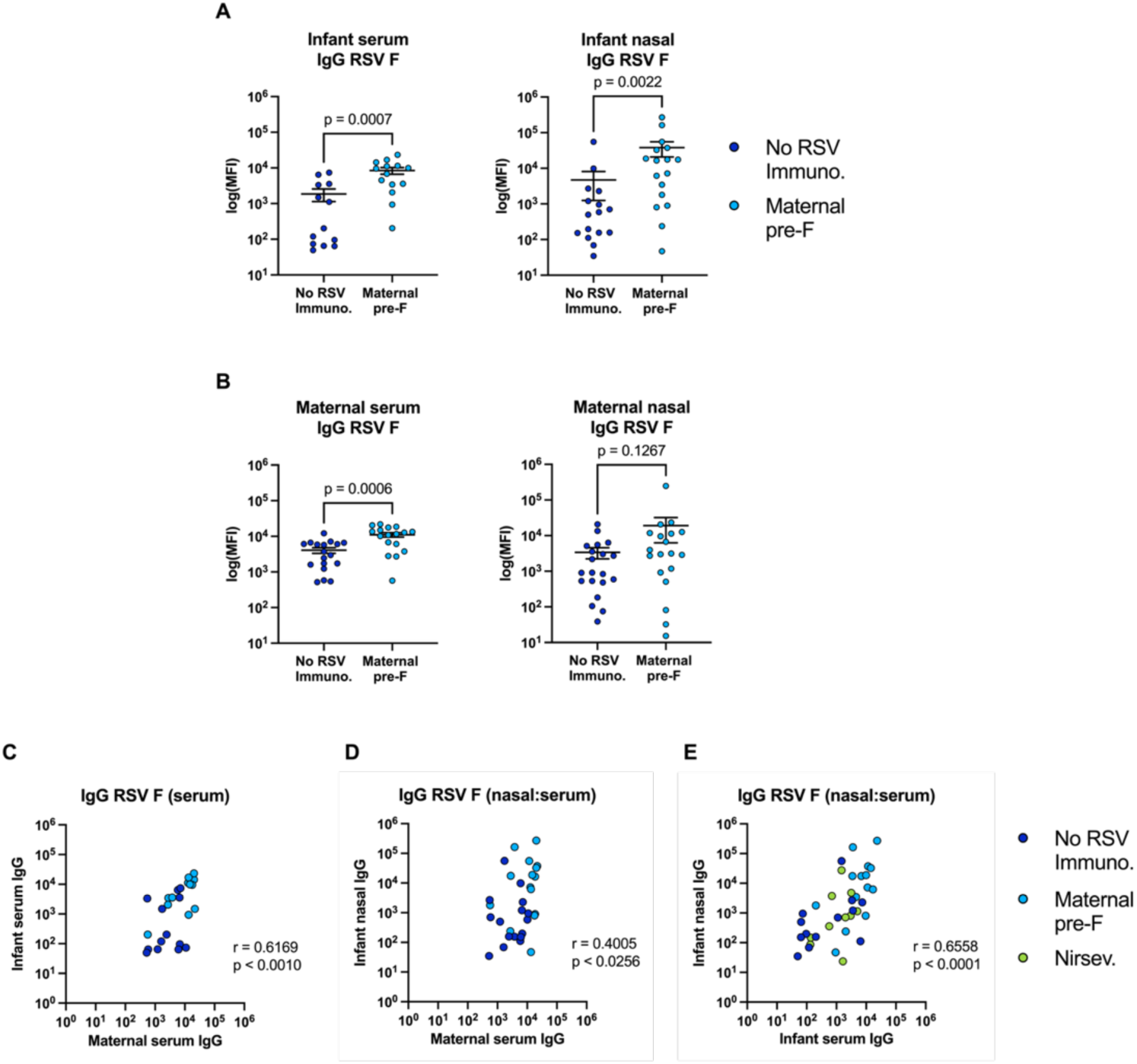
Maternal pre-F vaccination boosts infant serologic and mucosal immunity against RSV-A. Serum IgG levels against RSV-A fusion (F) protein in infants who have either received no RSV immunoprophylaxis (n=13) or whose mothers received the maternal pre-F vaccination (n=14) are shown, as well as nasal IgG RSV F protein levels in infants who have either received no RSV immunoprophylaxis (n=16) or whose mothers received the maternal pre-F vaccination (n=17) (**A**). Serum IgG F levels in mothers who received no RSV vaccine (n=18) or those who received the maternal pre-F vaccination (n=17), as well as nasal IgG F levels in mothers who received no RSV vaccine (n=20) or those who received the maternal pre-F vaccination (n=19), are also shown (**B**). Matched correlations between maternal serum IgG and infant serum IgG F levels (n=25) (**C**) and maternal serum IgG and infant nasal IgG (n=31) F levels (**D**) were performed. Overall correlation between matched infant nasal and serum IgG levels (n=37) in infants who received no RSV immunoprophylaxis, received nirsevimab, or those whose mothers received the maternal pre-F vaccination (**E**). Mean value and standard error of the mean are displayed. Two-tailed Mann-Whitney U tests were performed between individual groups. For the correlations, two-tailed Spearman correlations were performed between groups. Abbreviations: F, Fusion protein; Ig, immunoglobulin; MFI, median fluorescence intensity; Nirsev., Nirsevimab; No RSV Immuno, no RSV immunoprophylaxis.

### Higher maternal anti-RSV antibody titers drive higher infant airway mucosal antibody levels

To better understand how maternal anti-RSV antibody levels may correlate with infant airway mucosal antibody levels, we next analyzed blood and nasal swab samples collected from the mothers of infants in the study. Mothers were grouped into two categories: maternal pre-F (mothers received the maternal pre-F vaccine while pregnant), and no RSV immunoprophylaxis (mothers who did not receive the maternal pre-F vaccine). As expected, anti-RSV-F IgG titers in serum were significantly higher in mothers who received pre-F vaccination (**Figure 4B, left**, p = 0.0006). We did not detect a difference in nasal anti-F IgG titers in mothers (**Figure 4B, right**). Similar findings were seen for antibodies against RSV-B **(Supplemental Figure 3B).** This lack of difference may suggest that maternal nasal anti-RSV-F IgG levels are at least in part independent of serum levels and potentially more reflective of prior natural infection rather than vaccination in adults.

We then compared anti-RSV antibody levels between mothers and infants to assess for correlation related to transplacental transfer. There was strong correlation between maternal serum anti-RSV-F IgG and both infant serum and nasal anti-F antibodies (**Figure 4C**, p < 0.0010; **Figure 4D**, p < 0.0256; **Supplemental Figure 3C, 3D**). We also observed a significant correlation between anti-RSV-F IgG in the infant serum and their nasal mucosa (**Figure 4E**, p < 0.0001; **Supplemental Figure 3E**). These results suggest that strategies that boost serum anti-RSV IgG levels in infants translate into higher nasal anti-RSV antibody levels, which may play an important role in optimizing protection. As expected, we did not detect differences between maternal serum anti-RSV-A or anti-RSV-B IgA and infant serum or nasal IgA levels because IgA is not transplacentally transferred **(Supplemental Figures 5A,5B, 6A, 6B)**. Additionally, nasal and serum IgA are generated through distinct mechanisms, and thus we were not expecting to see a correlation between the two sources of IgA in infants **(Supplemental Figures 5E, 6E)**.

**Figure 5.**
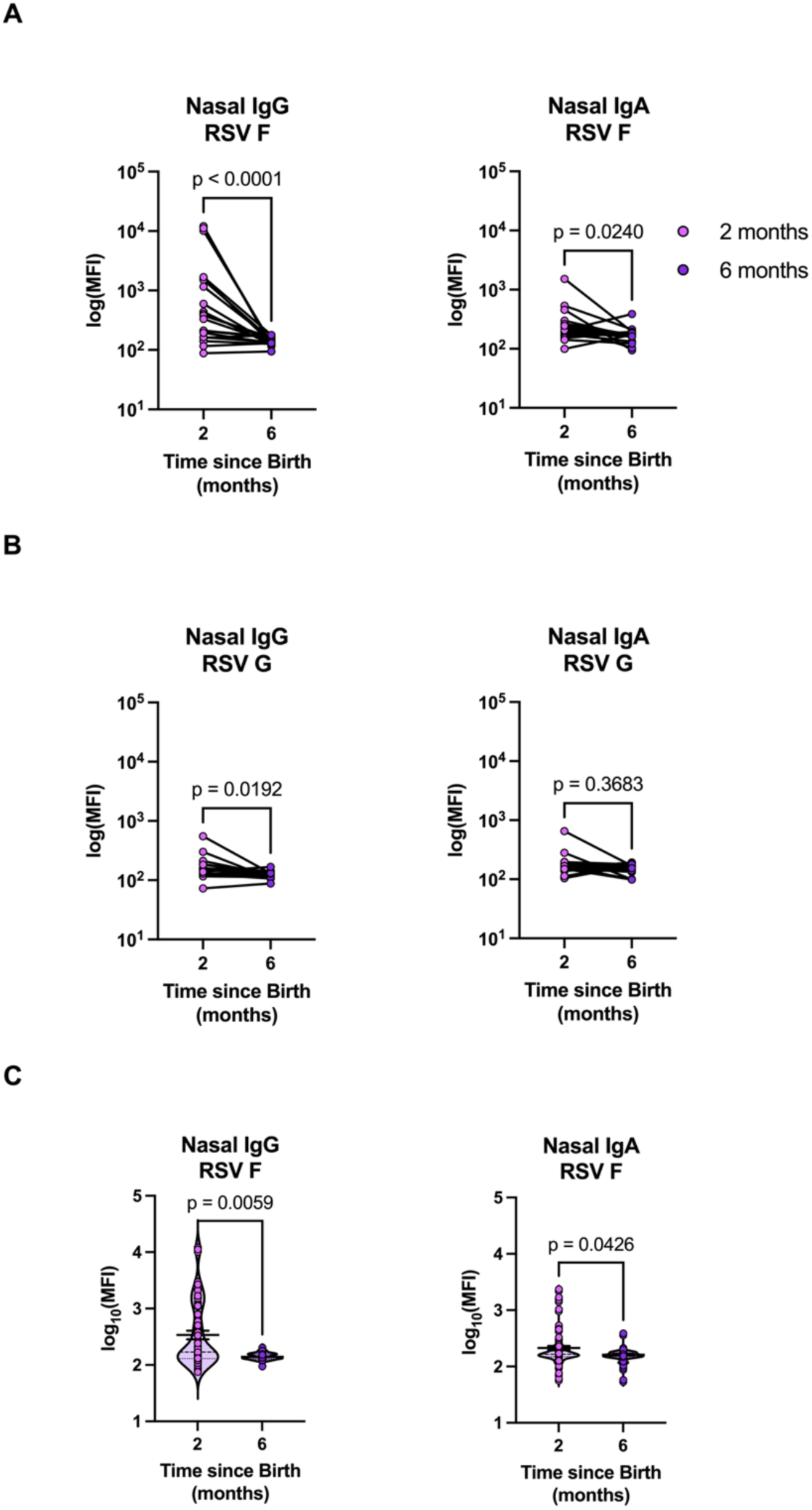
Durability of the infant mucosal response to RSV-A wanes with time following maternal vaccination. Matched nasal IgG (left) and IgA (right) levels against RSV-A fusion (F) protein (**A**) and attachment (G) protein (**B**) in infants two- and six-months following birth are shown (n=20). Violin plots representing log_10_ transformed nasal IgG (left) and IgA (right) F levels for all infants (**C**) are shown (n=67). The heavy dashed line indicates the median and the fine dashed lines indicate quartiles. Two-tailed Wilcoxon signed-rank tests were performed between the pairs. Abbreviations: F, Fusion protein; Ig, immunoglobulin; MFI, median fluorescence intensity.

### Nasal mucosal protection in infants from maternal pre-F vaccination wanes over time

To assess the durability of maternal pre-F-induced mucosal antibody responses in infants, we longitudinally analyzed nasal swab samples from infants whose mothers had received the maternal pre-F vaccine, collecting samples two- and six-months post birth (“Longitudinal Cohort”; **Figure 1A**). Sixty-seven infants enrolled at birth were included in the cohort, with 31 (46%) females (**Figure 1B**). Across matched samples from the twenty infants who had nasal samples collected at both two- and six-months post birth, we observed that nasal IgG titers against the RSV-F protein decreased significantly at six months of age compared to nasal titers at two months of age (**Figure 5A, left**, p < 0.0001). Interestingly, mean nasal IgG against the RSV-A G protein was elevated at 2 months of age, also waned significantly by 6 months (**Figure 5B, left**, p = 0.0192), suggesting that some mothers transferred anti-RSV-G antibodies transplacentally, most likely as the result of prior maternal natural infection. While it is possible that natural infection in the infants could have elicited an increase in nasal IgG against RSV G protein as well, infants were only included in this cohort if mothers reported the absence of respiratory illnesses in their infants before 6 months of age. Samples collected from the full 67-infant cohort, which includes an additional 47 nasal samples from infants who provided samples at only one time point, further illustrated the waning of anti-RSV-F IgG titers (**Figure 5C, left**, p = 0.0059). Similarly, IgG and IgA levels against the F and G proteins of RSV-B waned across the matched and total cohort **(Supplemental Figure 7)**.

A small subset of participants exhibited elevated nasal anti-RSV-F IgA levels at the two-month timepoint within the matched (**Figure 5A, right**, p = 0.0240) and unmatched groups **Figure 5C, right**, p = 0.0426), which may reflect natural infection or maternal transfer of secretory IgA, potentially through breastfeeding. As with nasal IgG, anti-RSV nasal IgA titers waned over 6 months of age, potentially corresponding with weaning from breastfeeding, although a connection between mucosal anti-RSV IgA and breastfeeding has not been well defined. Overall, these results indicate that nasal antibody protection against RSV wanes considerably within the first six months of life.

### Conserved nasal and serum responses across RSV strain and isotype

Because we were interested in further examining the relationships between anti-RSV antibody responses across isotypes and strains, we performed correlational analyses of both nasal and serum IgG and IgA levels against F and G proteins from infants in the RSV season cohort. Nasal IgG responses against RSV-A and RSV-B were significantly positively correlated for both fusion and G proteins, suggesting positive correlations among cross- strains and RSV surface glycoprotein types (**Figure 6A**). Similar positive correlations were seen for IgA. In the serum, IgG and IgA levels were not significantly correlated with one another, exemplifying that the immunological processes driving the production of each isotype within the nasal compartment might be distinct from that of the blood (**Figure 6B**).

**Figure 6.**
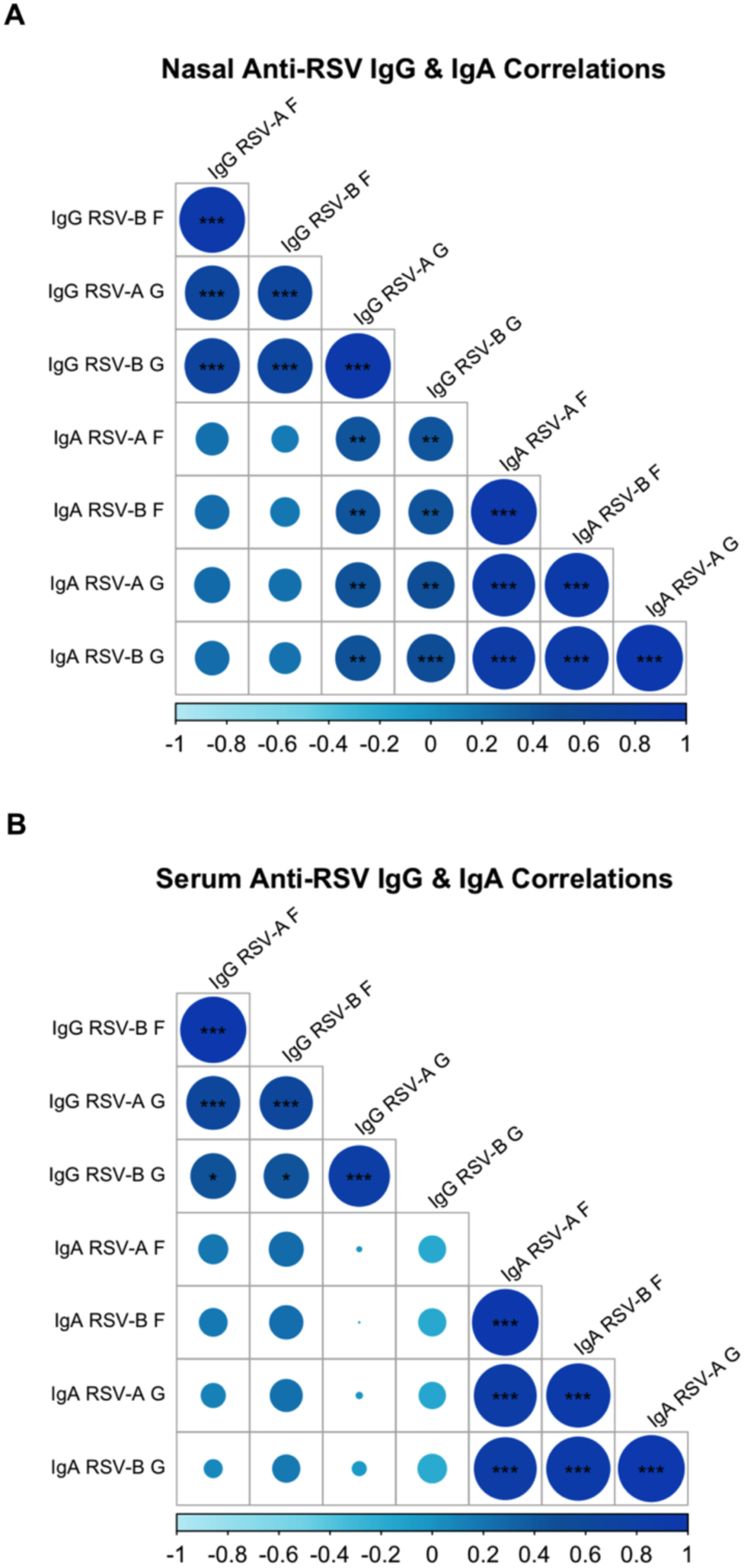
Spearman correlation matrices between nasal (n=44) (**A**) and serum (n=37) (**B**) IgG and IgA F and G levels against RSV-A or RSV-B fusion (F) protein or G (attachment) protein from infants in the RSV Season cohort. Analysis performed with Spearman correlation with Benjamini-Hochberg correction for multiple comparisons. Abbreviations: Ig, immunoglobulin.

## Discussion

Here, we examined airway mucosal antibody responses against RSV in infancy. Higher nasal anti-RSV antibody titers were associated with a lower likelihood of medically attended and clinically significant RSV. Infant nasal anti-RSV IgG closely correlated with maternal and infant serum IgG but waned over time, suggesting that passively acquired antibodies reach the airway surface and provide potentially meaningful yet finite protection.

RSV is first encountered at the airway mucosa, where local antibodies may limit infection or disease. In adults, low mucosal anti-RSV IgA is an independent risk factor for RSV infection.[9] Because IgA is not transferred transplacentally and is generally induced by infection, nasal anti-RSV IgA was very low in young infants but increased among those who had experienced at least one RSV season. In this older group, higher mucosal anti-RSV IgA appeared to be associated with a lower likelihood of medically attended RSV, suggesting a potential protective role similar to that observed in adults. However, natural infection cannot be relied upon to establish protection during early infancy, when the burden of severe RSV disease is greatest[2, 10], and strategies that optimize RSV immunity without relying on natural infection are essential.

In the absence of natural infection to induce IgA, mucosal IgG may be particularly important for protection against RSV. Prior studies demonstrated that mucosal, but not systemic, anti-RSV IgG correlated with lower viral load and reduced airway inflammation during RSV infection[11], although these studies preceded the availability of effective maternal RSV vaccination. Consistent with these observations, we found lower mucosal anti-RSV levels in infants with RSV infection. Nasal IgG correlated with both infants and maternal serum IgG, supporting a model in which maternally derived IgG is transferred transplacentally and subsequently transported across infant’s airway via the neonatal Fc receptor (FcRn). Our study further focused on antibodies targeting the RSV fusion protein, the target of current maternal RSV vaccines and a key mediator of protection. Anti-F but not anti-G appeared to be associated with lower likelihood of medically attended RSV during their first RSV season. The lack of association for anti-G antibodies may reflect their low abundance in early infancy, diminished cross reactivity[12], and tendency to wane rapidly[13]. Together, these findings suggest that maximizing maternal transfer and persistence of anti-F IgG may strengthen mucosal protection in early infancy, although larger longitudinal studies are needed to define the magnitude and durability of this protection.

While the gestational timing of maternal vaccination and the infant’s birth relative to the RSV season may influence the magnitude and duration of transferred antibody[13], breastfeeding may also play an important role in protection at the surface of the upper airway. As infants breastfeed, secretory IgA (sIgA) and IgG are transferred from mother to infant[14], coating the naso/oropharynx and providing transient immunity. Antibodies from breastmilk provide short-lived protection[15, 16], thus immunity from breastmilk requires ongoing exposure. Additional studies are needed to determine the impact of maternal vaccination with pre-F vaccine, its impact on breastmilk, and the protective potential in infants.

Our findings are consistent with others that suggest sufficient mucosal RSV antibodies, especially mucosal IgA against Pre-F[17],are important for protection against RSV bronchiolitis[18]. While our study overcomes the limits of a small pediatric cohort in prior studies, our study has several limitations. Although nearly 300 infants were included overall and findings were consistent across cohorts, each cohort was modest, and the observational design limits causal inference. The study also lacked the dense longitudinal sampling needed to characterize individual antibody trajectories or fully compare immunization strategies and breastmilk-derived immunity. Larger prospective studies incorporating serial maternal and infant serum, nasal, and breastmilk samples together with clinical outcomes are needed to define the level and duration of mucosal antibody associated with protection.

In conclusion, higher nasal anti-RSV antibody levels were associated with a lower likelihood of medically attended RSV, and maternal vaccination and infant monoclonal antibody administration were associated with higher systemic and mucosal antibody levels. However, passively acquired mucosal antibodies waned over time, emphasizing the finite duration of passive immunity. Strategies that increase the magnitude and durability of airway antibodies may further reduce RSV morbidity during early infancy.

## Methods

### Emergency Department Cohort Participant Recruitment and Enrollment

Children 6 years of age and under presenting to the Emergency Department at Boston Children’s Hospital (BCH) who were undergoing a clinically obtained nasopharyngeal swab respiratory viral panel were enrolled under Institutional Review Board number (IRB) P00028229. Written informed consent was acquired from parents/legal guardians. All clinical and demographic data was entered into the REDCap database.

### RSV Season Cohort and Longitudinal Cohort Participant Recruitment and Enrollment

Mothers and their infants aged 18 months or younger seen in primary care clinic, inpatient units, and obstetric clinics at Massachusetts General Hospital were approached between November 2024 and April 2025 (RSV Season cohort) and between February 2024 and January 2026 (Longitudinal cohort) for enrollment under IRB number 2020P0000955.

Potential participants were identified via clinic schedules, referrals from practitioners, or inpatient lists. Eligible mothers were at least 18 years old and able to provide written informed consent for sample collection for both themselves and their infants. Vaccination status and demographic information were obtained from electronic medical records.

### RSV Season and Longitudinal Cohort Sample Collection and Processing

Serum and nasal swab samples were collected during the enrollment visit from mothers and infants and from infants during subsequent follow-up outpatient appointments. Blood was collected using a microneedle capillary blood collection device (TAP II blood collection device, YourBio Health Devices). Blood was centrifuged at 2500 g for 5 minutes at room temperature, after which the serum was aliquoted into cryogenic vials and stored at -80°C until antibody analysis. Nasal swab samples were collected by study staff using FLOQSwabs (Copan flocked swabs) following the manufacturer’s instructions. One anterior nasal swab was used to swab both nostrils of each participant before being placed into a 15 mL conical tube containing 3 mL of phosphate buffer solution (PBS). These conical tubes containing swabs were then transported from clinical areas to the laboratory for processing. Processing of the anterior nasal swab entailed discarding the swab and aliquoting the 3 mL of PBS into three cryogenic vials, which were stored at -80°C until use.

### RSV viral testing

Following completion of clinically indicated viral testing employing a nasopharyngeal swab in the Emergency Department, discarded viral transport medium (VTM) was retrieved and stored at -80°C until use. In this study, VTM samples collected from September to April of 2023-24, 2024-25 and 2025-26 were included and analyzed. Lab-based RT-PCR tests for RSV A and RSV B were performed using the VTM samples. Briefly, viral RNA was extracted using QIAamp MinElute Virus Spin Kit (QIAGEN) following manufacturer’s instructions.

One-step RT-PCR was performed using TaqMan^TM^ assay RSV-A (Vi99990014_po) and RSV-B (Vi99990015_po) to confirm the viral infection status.

### RSV Specific Antibody Detection

RSV-A prefusion (pre-F) protein and RSV-A postfusion (post-F) protein were purchased from Acro biosystems. RSV-B fusion (F) protein, RSV-A glycoprotein (G) and RSV-B glycoprotein (G) were purchased from Sino biological. RSV proteins were covalently linked to different Luminex beads (Bio-Plex Pro Magnetic COOH Beads, BIO-RAD) with 6µg of proteins for 1.25x10^6^ beads using the Bio-Plex Amine Coupling Kit (BIO-RAD), following manufacturer’s instructions. Control beads for total IgG and IgA were purchased from Millipore Sigma.

Serum samples were diluted to 1:1000, and nasal samples were thawed and centrifuged at 3000xg for 10 mins before use without further dilution. 50uL of samples (diluted serum of undiluted nasal samples) were then added to antigen-coupled magnetic Luminex bead mix to form immune complexes. After an incubation for 2hrs at room temperature, shaking at 850 rpm, secondary antibody PE-conjugated mouse anti-human IgG1 or IgA1 was added at 1.3µg/mL, and then incubated for 1.5hrs at room temperature, shaking at 850 rpm. A fixation step with 4% PFA was included following the final washes for safety. Median fluorescence intensity (MFI) was measured using the Luminex^TM^ 200^TM^ system. A serum sample from an adult donor who recovered from recent confirmed RSV infection was included in every plate and served as positive control. Normal rat serum (1:1000 diluted) was used as negative control. Total IgG or IgA control beads were used for normalization.

### Statistical analysis

Statistical analyses were performed using GraphPad Prism version 11.0.2. Comparisons between two independent groups were conducted using the Mann-Whitney U test, whereas paired comparisons were performed using the Wilcoxon matched-pairs signed-rank test. Associations between continuous variables were assessed using Spearman’s rank correlation. Correlation plots were generated using R version 2026.01.0+392, using packages ‘psych,’ ‘corrplot,’ ‘Hmisc’, and ‘readxl.’

A univariable logistic regression model and multivariable logistic regression model was fitted to evaluate their independent associations with RSV positivity. Results were reported as odds ratios (OR) with 95 % confidence intervals.

## Data Availability

All data produced in the present study are available upon reasonable request to the authors

## Acknowledgements

This work was funded by NIH/NHLBI: R01HL173059 (LY); NIH/NIAID: 1U19AI167899 (AGE), MGH ECOR: MGH Research Scholar Award (AGE), Massachusetts Consortium on Pathogen Readiness Award (KES), Massachusetts Consortium on Pathogen Readiness Award (LMY and PPM), and the Chan-Zuckerberg Initiative (LY, WIG, BHH).

## Conflicts

AGE has received research funding from Merck Pharmaceuticals; this relationship was reviewed and managed by Massachusetts General Hospital and is in accordance with the MGH conflict of interest policies. The remaining authors have disclosed that they do not have any potential conflicts of interest.

---

**Supplemental Figure 1.**
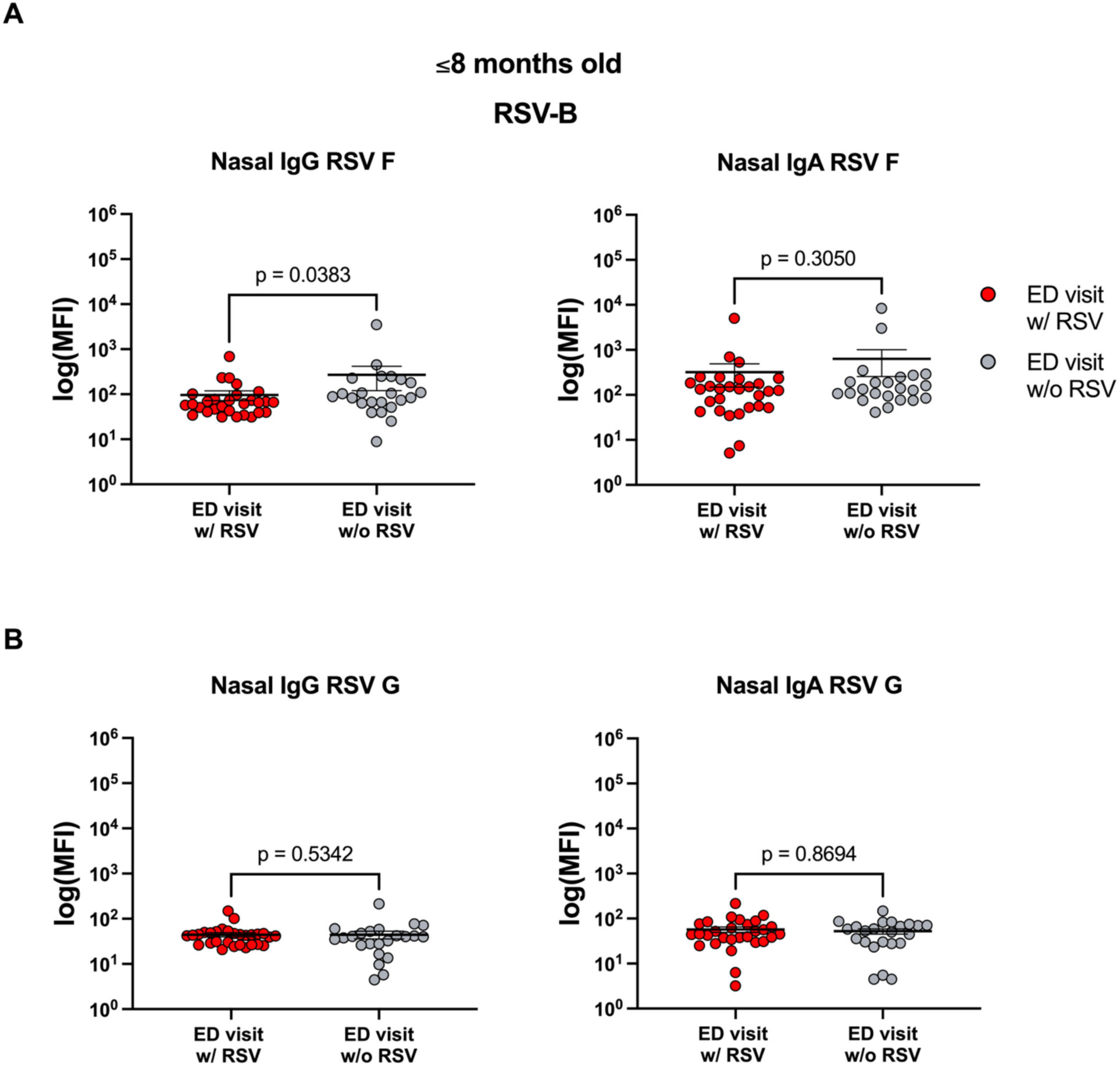
Nasal antibodies against RSV-B in infants ≤8 months of age. Nasal (left) IgG and IgA (right) levels against RSV-B fusion (F) protein (**A**) and attachment (G) protein (**B**) in infants ≤8 months of age with RSV (n=29) and without RSV (n=23) are shown. Mean value and standard error of the mean are displayed. Two-tailed Mann-Whitney U tests were performed between individual groups. Abbreviations: Ig, immunoglobulin; MFI, median fluorescence intensity.

**Supplemental Figure 2.**
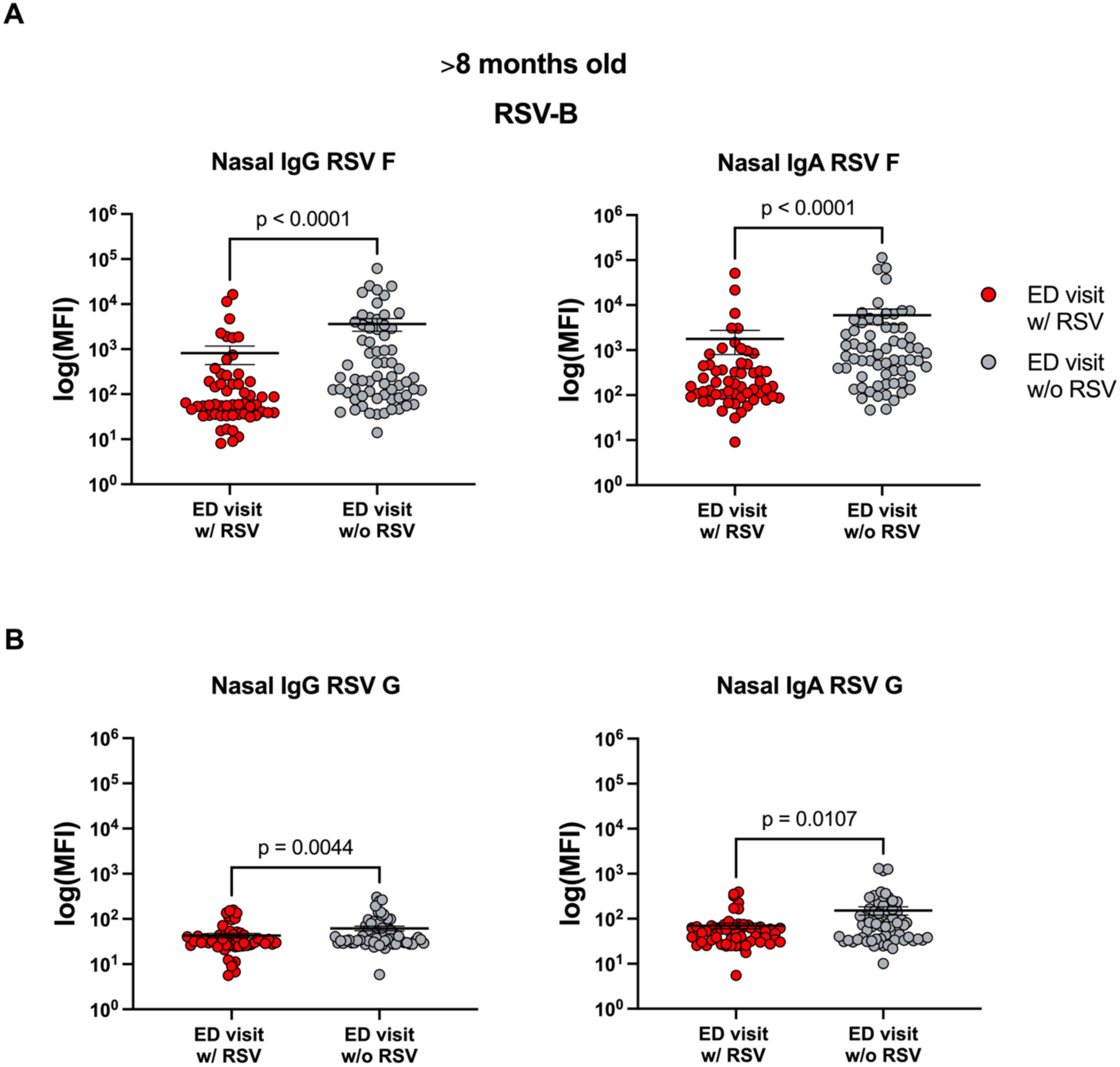
Nasal antibodies against RSV-B in infants older than 8 months of age. Nasal (left) and IgA (right) levels against RSV-B fusion (F) protein (**A**) and attachment (G) protein (**B**) in children >8 months of age with RSV (n=56) and without RSV (n=66) are shown. Mean value and standard error of the mean are displayed. Two-tailed Mann-Whitney U tests were performed between individual groups. Abbreviations: Ig, immunoglobulin; MFI, median fluorescence intensity.

**Supplemental Figure 3.**
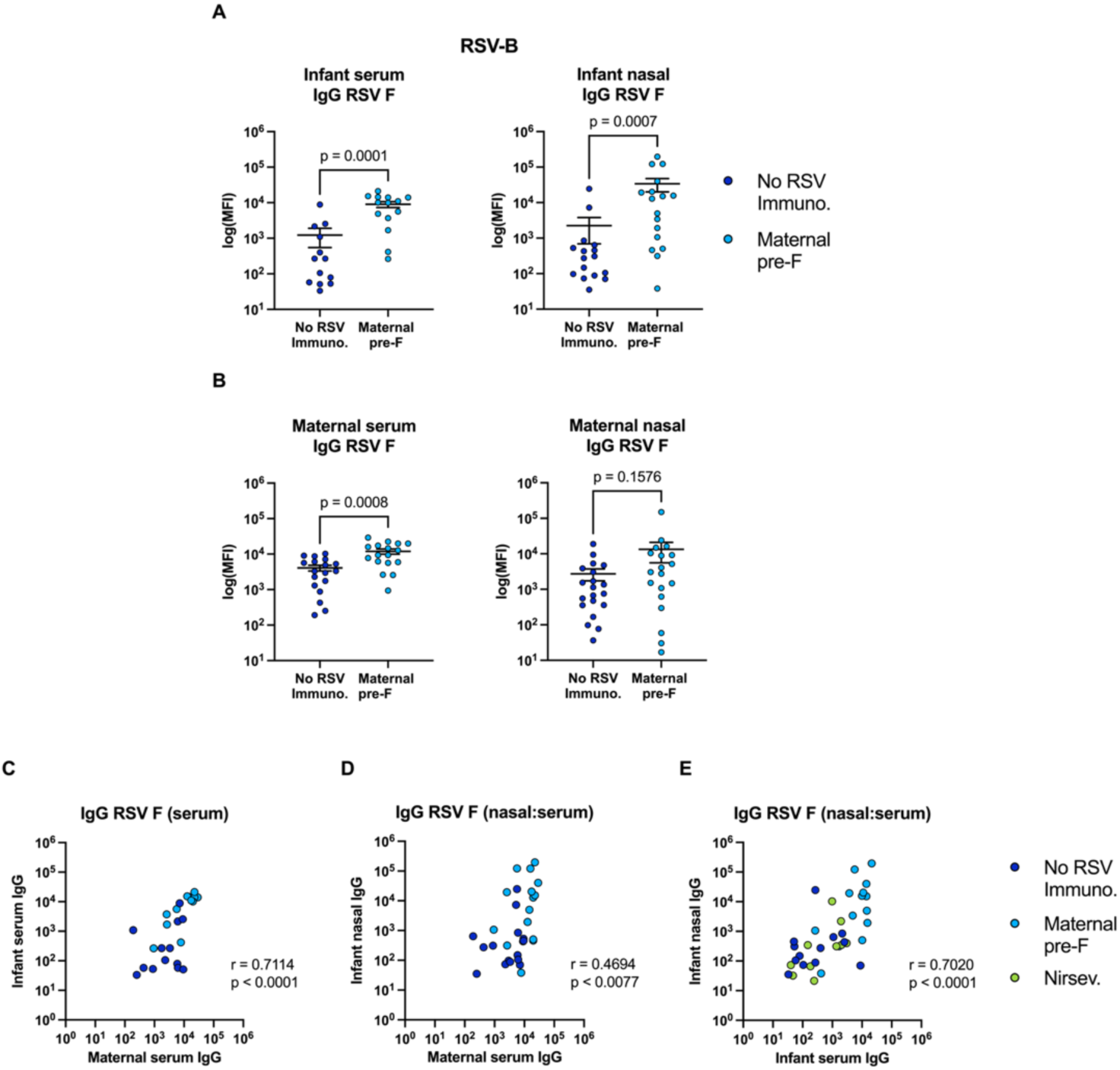
Maternal pre-F vaccination boosts infant serologic and mucosal immunity against RSV-B. Serum IgG levels against RSV-B fusion (F) protein in infants who have either received no RSV immunoprophylaxis (n=13) or whose mothers received the maternal pre-F vaccination (n=14) are shown, as well as nasal IgG F levels in infants who have either received no RSV immunoprophylaxis (n=16) or whose mothers received the maternal pre-F vaccination (n=17) (**A**). Serum IgG F levels in mothers who received no RSV vaccine (n=18) or those who received the maternal pre-F vaccination (n=17), as well as nasal IgG F levels in mothers who received no RSV vaccine (n=20) or those who received the maternal pre-F vaccination (n=19), are also shown (**B**). Correlations between matched maternal serum IgG and infant serum IgG levels (n=25) (**C**) and maternal serum IgG and infant nasal IgG F levels (n=31) (**D**) were performed. Overall correlation between matched infant nasal and serum IgG levels (n=37) in infants who received no RSV immunoprophylaxis, received nirsevimab, or those whose mothers received the maternal pre-F vaccination (**E**). Mean value and standard error of the mean are displayed. Two-tailed Mann-Whitney U tests were performed between individual groups. For the correlations, two-tailed Spearman correlations were performed between groups. Abbreviations: F, Fusion protein; Ig, immunoglobulin; MFI, median fluorescence intensity; Nirsev., Nirsevimab; No RSV Immuno, no RSV immunoprophylaxis.

**Supplemental Figure 4.**
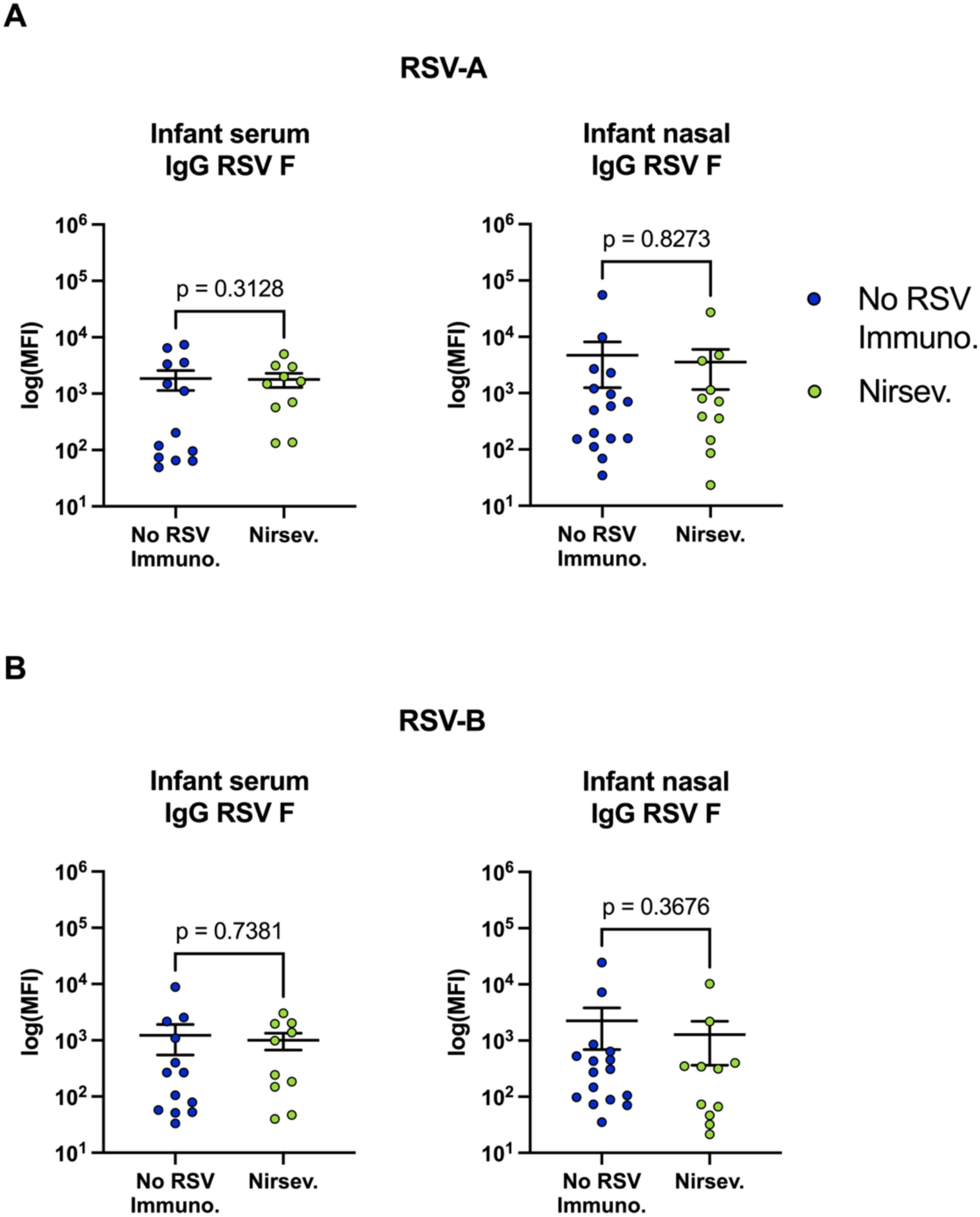
No significant difference is shown in immune responses between infants who received no RSV immunoprophylaxis and those who received nirsevimab. Serum IgG levels against RSV-A fusion (F) protein (**A**) or RSV-B F protein (**B**) in infants who have either received no RSV immunoprophylaxis (n=13) or nirsevimab (n=10) are shown, as well as nasal IgG F levels in infants who have either received no RSV immunoprophylaxis (n=16) or nirsevimab (n=11). Two-tailed Mann-Whitney U tests were performed between individual groups. Abbreviations: Ig, immunoglobulin; MFI, median fluorescence intensity; Nirsev., nirsevimab; No RSV Immuno, no RSV immunoprophylaxis.

**Supplemental Figure 5.**
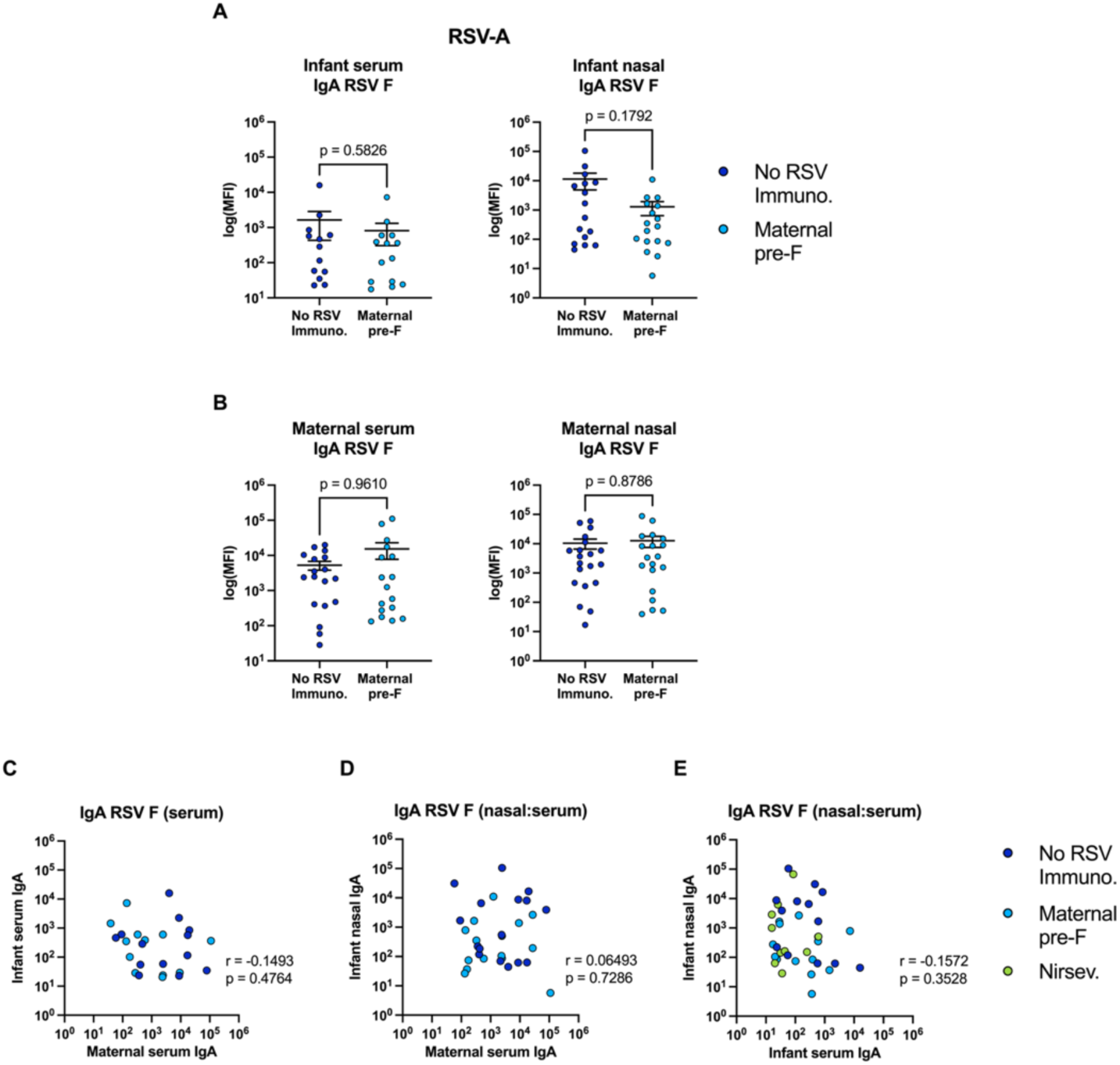
No significant difference is shown in anti-RSV-A IgA titers between infants who received no RSV immunoprophylaxis and those who received nirsevimab. Serum IgA levels against RSV-A fusion (F) protein in infants who have either received no RSV immunoprophylaxis (n=13) or whose mothers received the maternal pre-F vaccination (n=14) are shown, as well as nasal IgA F levels in infants who have either received no RSV immunoprophylaxis (n=16) or whose mothers received the maternal pre-F vaccination (n=17) (**A**). Serum IgA F levels in mothers who received no RSV vaccine (n=18) or those who received the maternal pre-F vaccination (n=17), as well as nasal IgA F levels in mothers who received no RSV vaccine (n=20) or those who received the maternal pre-F vaccination (n=19), are also shown (**B**). Correlations between matched maternal serum IgA and infant serum IgA levels (n=25) (**C**) and maternal serum IgA and infant nasal IgA F levels (n=31) (**D**) were performed. Overall correlation between matched infant nasal and serum IgA levels (n=37) in infants who received no RSV immunoprophylaxis, received nirsevimab, or those whose mothers received the maternal pre-F vaccination (**E**). Mean value and standard error of the mean are displayed. Two-tailed Mann-Whitney U tests were performed between individual groups. For the correlations, two-tailed Spearman correlations were performed between groups. Abbreviations: F, Fusion protein; Ig, immunoglobulin; MFI, median fluorescence intensity; Nirsev, Nirsevimab; No RSV Immuno, no RSV immunoprophylaxis.

**Supplemental Figure 6.**
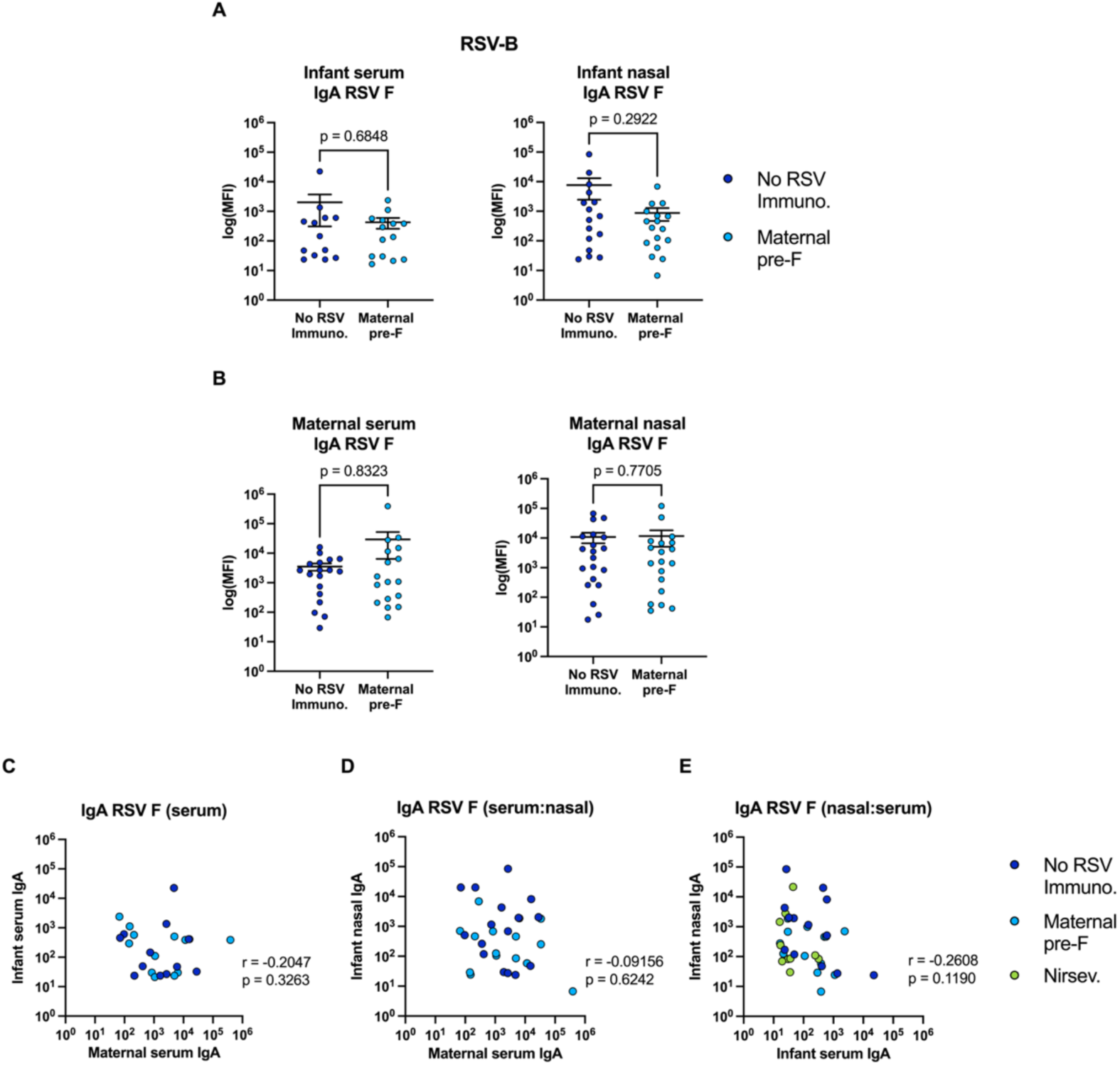
No significant difference is shown in anti-RSV-B IgA titers between infants who received no RSV immunoprophylaxis and those who received nirsevimab. Serum IgA levels against RSV-B fusion (F) protein in infants who have either received no RSV immunoprophylaxis (n=13) or whose mothers received the maternal pre-F vaccination (n=14) are shown, as well as nasal IgA F levels in infants who have either received no RSV immunoprophylaxis (n=16) or whose mothers received the maternal pre-F vaccination (n=17) (**A**). Serum IgA F levels in mothers who received no RSV vaccine (n=18) or those who received the maternal pre-F vaccination (n=17), as well as nasal IgA F levels in mothers who received no RSV vaccine (n=20) or those who received the maternal pre-F vaccination (n=19), are also shown (**B**). Correlations between matched maternal serum IgA and infant serum IgA levels (n=25) (**C**) and maternal serum IgA and infant nasal IgA F levels (n=31) (**D**) were performed. Overall correlation between matched infant nasal and serum IgA levels (n=37) in infants who received no RSV immunoprophylaxis, received nirsevimab, or those whose mothers received the maternal pre-F vaccination (**E**). Mean value and standard error of the mean are displayed. Two-tailed Mann-Whitney U tests were performed between individual groups. For the correlations, two-tailed Spearman correlations were performed between groups.

**Supplemental Figure 7.**
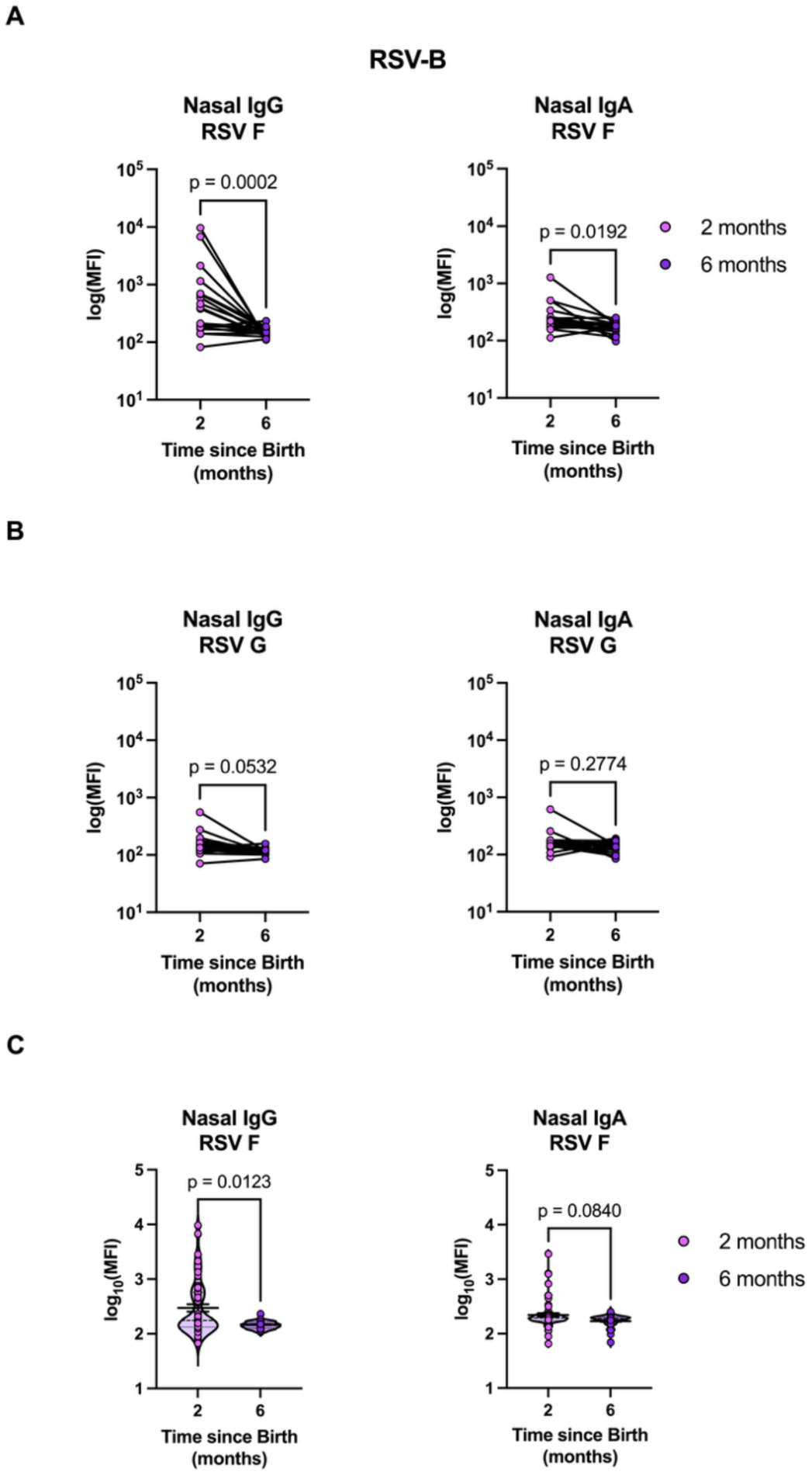
The infant mucosal response to RSV-B dampens with time following maternal vaccination. Matched nasal anti-RSV-B IgG and IgA levels against the fusion (F) protein (**A**) and attachment (G) protein (**B**) in infants two- and six-months following birth are shown (n=20). Two-tailed Wilcoxon signed-rank tests were performed between the timepoints. Violin plots representing log_10_transformed nasal IgG (left) and IgA (right) levels against RSV-B fusion (F) protein (**C**) for all infants two- and six-months following birth are shown (n=67). The heavy dashed line indicates the median and the fine dashed lines indicate quartiles. Two-tailed Mann-Whitney U tests were performed between individual groups. Abbreviations: Ig, immunoglobulin; MFI, median fluorescence intensity.

## Notes

### Author Declarations

Both Massachusetts General Hospital and Boston Children's Hospital Institutional Review Boards gave ethical approval for this work under the following protocols: MGH IRB number 2020P0000955 and BCH IRB number P00028229.

## References

1. Coindy EL, Efstathiou C, Talwar S, et al. Antibody-mediated protection against respiratory syncytial virus in children. Eur Respir Rev 2024; 33.

2. Li Y, Wang X, Blau DM, et al. Global, regional, and national disease burden estimates of acute lower respiratory infections due to respiratory syncytial virus in children younger than 5 years in 2019: a systematic analysis. Lancet 2022; 399:2047–64.

3. Greenough A, Cox S, Alexander J, et al. Health care utilisation of infants with chronic lung disease, related to hospitalisation for RSV infection. Arch Dis Child 2001; 85:463–8.

4. Sarna M, Ware RS, Lambert SB, Sloots TP, Nissen MD, Grimwood K. Timing of First Respiratory Virus Detections in Infants: A Community-Based Birth Cohort Study. J Infect Dis 2018; 217:418–27.

5. Hammitt LL, Dagan R, Yuan Y, et al. Nirsevimab for Prevention of RSV in Healthy Late-Preterm and Term Infants. N Engl J Med 2022; 386:837–46.

6. Kampmann B, Madhi SA, Munjal I, et al. Bivalent Prefusion F Vaccine in Pregnancy to Prevent RSV Illness in Infants. N Engl J Med 2023; 388:1451–64.

7. Silk BJ, Prill MM, Winn AK, et al. Respiratory Virus Activity - United States, July 1, 2024- June 30, 2025. MMWR Morb Mortal Wkly Rep 2026; 75:77–84.

8. Tang Y, Boribong BP, Swank ZN, et al. COVID-19 mRNA Vaccines Induce Robust Levels of IgG but Limited Amounts of IgA Within the Oronasopharynx of Young Children. J Infect Dis 2024; 230:1390–9.

9. Walsh EE, Falsey AR. Humoral and mucosal immunity in protection from natural respiratory syncytial virus infection in adults. J Infect Dis 2004; 190:373–8.

10. Hall CB, Weinberg GA, Blumkin AK, et al. Respiratory syncytial virus-associated hospitalizations among children less than 24 months of age. Pediatrics 2013; 132:e341–8.

11. Vissers M, Ahout IM, de Jonge MI, Ferwerda G. Mucosal IgG Levels Correlate Better with Respiratory Syncytial Virus Load and Inflammation than Plasma IgG Levels. Clin Vaccine Immunol 2015; 23:243–5.

12. Ogunsemowo O, Olaleye DO, Odaibo GN. Genetic diversity of human respiratory syncytial virus circulating among children in Ibadan, Nigeria. PLoS One 2018; 13:e0191494.

13. Jasset OJ, Lopez Zapana PA, Bahadir Z, et al. Enhanced placental antibody transfer efficiency with longer interval between maternal respiratory syncytial virus vaccination and birth. Am J Obstet Gynecol 2025; 232:554 e1–e15.

14. Atyeo C, Alter G. The multifaceted roles of breast milk antibodies. Cell 2021; 184:1486–99.

15. Aldred MJ, Vaughan AG, Yuill SJ, Wade WG, Walker DM. Persistence of IgA in neonatal saliva following breast feeding. Early Hum Dev 1986; 14:273–6.

16. Yang X, Fox A, DeCarlo C, Pineda N, Powell RLR. The Secretory IgA Response in Human Milk Against the SARS-CoV-2 Spike Is Highly Durable and Neutralizing for At Least 1 Year of Lactation Postinfection. Breastfeed Med 2023; 18:602–11.

17. Hodgson D, Jarju S, Gatcombe L, et al. Mucosal IgA to pre-fusion F protein predicts protection from RSV infection in a high burden setting. Nat Commun 2026; 17.

18. Pillet S, Cantais A, Noailly B, et al. Mucosal nirsevimab levels in respiratory syncytial virus breakthrough bronchiolitis. Lancet Infect Dis 2024; 24:1192–4.

